# Clinical diagnosis of COVID-19: a prompt, feasible, and sensitive diagnostic tool for COVID-19 based on a 1,757-patient cohort (The AndroCoV Clinical Scoring for COVID-19 diagnosis)

**DOI:** 10.1101/2020.12.23.20248803

**Authors:** Flávio Adsuara Cadegiani, John McCoy, Carlos Gustavo Wambier, Andy Goren

## Abstract

**Importance:** In the COVID-19 pandemic, a limiting barrier for more successful approaches to COVID-19 is the lack of appropriate timing for its diagnosis, during the viral replication stage, when antiviral approaches could demonstrate efficacy, precluding progression to severe stages. Three major reasons that hamper the diagnosis earlier in the disease are the unspecific and mild symptoms in the first stage, the cost- and time-limitations of the rtPCR-SARS-CoV-2, and the insufficient sensitivity of this test as desired for screening purposes during the pandemic. More sensitive and earlier methods of COVID-19 detection should be considered as key for breakthrough changes in the disease course and response to specific therapeutic strategies. Our objective was to propose a clinical scoring for the diagnosis of COVID-19 (The AndroCoV Clinical Scoring for COVID-19 Diagnosis) that has been validated in a large population sample, aiming to encourage the management of patients with high pre-clinical likelihood of presenting COVID-19, at least during the pandemics, independent of a rtPCR-SARS-COV-2 test.

**Materials and methods:** This is a compounded retrospective and prospective analysis of clinical data prospectively collected from the Pre-AndroCoV and AndroCov Trials that resulted in a clinical scoring for COVID-19 diagnosis based on likelihood of presenting COVID-19 according to the number of symptoms, presence of anosmia, and known positive household contact, in a variety of combinations of scoring criteria, aiming to the detect scorings that provided the highest pre-test probability and accuracy. Sensitivity, specificity, positive predictive value, negative predictive value, positive likelihood ratio, and accuracy were calculated for subjects screened in two different periods and altogether, for females, males, and both, in a total of nine different scenarios, for combinations between one, two, or three or more symptoms, or presence of anosmia in subjects without known positive household contacts, and no symptoms, one, two, or three or more symptoms, or presence of anosmia or ageusia in subjects with known positive household contacts.

**Results:** 1,757 patients were screened for COVID-19. Among the multiple combinations, requiring two or more symptoms with or without anosmia or ageusia for subjects without known contact and one or more symptoms with or without anosmia or ageusia with known positive contacts presented the highest accuracy (80.4%), and higher pretest probability and accuracy than virtually all rtPCR-SARS-CoV-2 commercially available kit tests.

**Conclusion:** The AndroCoV Clinical Scoring for COVID-19 Diagnosis was demonstrated to be a feasible, quick, inexpensive and sensitive diagnostic tool for clinical diagnosis of COVID-19. A clinical diagnosis of COVID-19 should avoid delays and missed diagnosis, and reduce costs, and should therefore be recommended as a first-line option for COVID-19 diagnosis for public health policies, at least while SARS-CoV-2 is the prevailing circulating virus.

**Key Points:** *Question:* Is a clinical diagnosis of COVID-19 sensitive, accurate, and feasible?

*Findings:* The present analysis of a 1,757-subject cohort of the AndroCoV trials demonstrated that clinical scoring for COVID-19 diagnosis can deliver a more sensitive and prompter diagnosis than the current gold-standard diagnostic method, rtPCR-SARS-CoV-2, with an accuracy above 80%.

*Meaning:* A clinical diagnosis of COVID-19 avoids missed diagnosis due to insufficient sensitivity or incorrect timing of the performance of rtPCR-SARS-CoV-2, reduces costs, avoid delays on specific managements, and allows the testing of potentially effective antiviral therapeutic approaches that should work if administered in the early stage of COVID-19

## Introduction

While COVID-19 pandemic has affected millions of people worldwide, its early stage remains poorly characterized (1). The inability to better understand the COVID-19 pathophysiology, clinical and biochemical presentation in the first days after contamination may be explained by a variety of challenging reasons. First, because symptoms in the first stage of COVID-19 are essentially unspecific, since it can resemble upper respiratory tract infection (URTI), dengue fever, and/or gastrointestinal (GI) infections (2), precluding subjects to be suspected for COVID-19 before the progression to more severe states, unless if they had known contact with a confirmed COVID-19 case. Second, because research on COVID-19 has mainly focused on approaches to reduce mortality in already severely affected COVID-19 subjects (1). Third, because even after the extensively described pathophysiology of second stage of COVID-19 as being basically mediated by overreactive, dysfunctional inflammatory responses (1), while virological activity becomes minor and not as relevant for the current clinical status, antiviral pharmacological approaches have been persistently and solely tested for this stage, when efficacy would not expect to be found. Without apparently effective approaches to early COVID-19, this stage has progressively been deprioritized in basic and clinical research.

A fourth reason that reinforces the unsuccessfulness of approaching early COVID-19 is that while COVID-19 is that while many are under-suspected due to lack of typical clinical characteristics, whenever COVID-19 is suspected, the need of a positive real time Polymerase Chain Reaction (rtPCR) for SARS-CoV-2 for the conclusive diagnosis of COVID-19, which remains as the gold standard diagnostic test for COVID-19, delays the time-to-diagnosis and time-to-treat. In addition, the sensitivity of the rtPCR-SARS-CoV-2 19 has demonstrated wide variability between kit tests (3-5), and may lead to an overwhelming number of false negative tests (6-12), which is particularly relevant for higher risk patients, allowing progression to severe states due to the inability to detect COVID-19 in earlier stages. Both increased time-to-diagnosis and false negative tests preclude patients from the correct timing of specific antiviral approaches for COVID-19, and may also have contributed to the lack of randomized clinical trials (RCTs) conducted during actual early COVID-19.

Hence, more sensitive and earlier detection of COVID-19 could be the key for a breakthrough change in the disease course and response to specific therapeutic strategies, since the majority of new molecules and drug repurposing focused on their potential antiviral activity, which would find the most effective results earliest in the disease.

Considering that: 1. Clinical or radiological criteria for other viral infections is the gold standard or a sufficient method for the diagnosis; 2. The need of a positive rtPCR-SARS-CoV-2 for the diagnosis of COVID-19 is a barrier in terms of cost and diagnostic delays; 3. Infections caused by other agents are unlikely to occur during the pandemic, when SARS-CoV-2 is the prevailing virus circulating and other infections are effectively prevented by the spread use of masks; 4. Since SARS-CoV-2 is the prevailing virus during the pandemics, a range of different and unspecific symptoms are more likely to be caused by this virus; and 5. For screening purposes, more sensitive tools than rtPCT-SARS-CoV-2 are highly recommended, our objective was to propose a clinical scoring for the diagnosis of COVID-19 that has been validated in a large population sample to encourage the management of patients with high pre-clinical likelihood of presenting COVID-19, at least during the pandemics, independently of the rtPCR-SARS-COV-2 result.

## Materials and methods

This is a retrospective analysis followed by a prospective analysis of clinical data that was prospectively collected from the Pre-AndroCoV and AndroCov Trials (13-16), as well as patients that followed up without participating or receiving any treatment regimen for COVID-19 aimed to calculate their likelihood to present COVID-19 according to the number of symptoms and contact with a known positive household.

Subjects presenting at least one of the following symptoms, that were actively searched, whether they had contact with confirmed case for COVID-19 or not, were screened for COVID-19 through a rtPCR-SARS-CoV-2, and included in the present analysis: 1 Specific COVID-19 manifestations: hyposmia, anosmia, dysgeusia or ageusia; 2. Symptoms typically present in dengue fever (dengue fever-like syndrome): myalgia, arthralgia, upper back pain, conjunctival hyperemia, pre-orbital pain; 3. Symptoms of upper respiratory tract infection (URTI) (URTI-like syndrome): nasal congestion, rhinorrhea, dry cough, self-reported perception of “sinusitis”, or self-reported perception of “sore throat”; 4. Symptoms of acute gastroenteritis (GE) (GE-like syndrome): nauseas, vomiting, or abdominal pain 5. Additional unspecific presentation, including lower back pain, leg pain, feverish, fatigue, weakness, dizziness and headache. For pre-existing symptoms or those that are frequently experimented, changes in the patterns of these symptoms were required in order to be counted as a symptom.

After the evaluation of the first 1,557 patients, 200 were presumedly diagnosed for COVID-19 based on the resulting clinical scoring when pre-test probability was higher than rtPCR-SARS-CoV-2 sensitivity, and prospectively evaluated. All 200 patients underwent a first rtPCR-SARS-CoV-2, and those with negative results underwent a second rtPCR-SARS-CoV-2, between 24 and 72 hours later the first one.

Combination of scenarios for clinical diagnosis of COVID-19 were tested for precision-related statistical parameters: when one, two, three or more symptoms, or when anosmia or ageusia were presented, and whether there was or there was not known positive households. Scenarios were tested for three moments, including two distinct periods and these two periods together. The first period comprised the observational study of the AndroCoV Trial (pre-AndroCoV Trial), between May 2020 and July 2020, and the second period, that comprised the AndroCoV RCTs and the follow-up of untreated patients that did not participate in any of the RCTs, between July 2020 to December 2020. Each scenario in each moment was analyzed for males, females, and overall.

### Statistical analysis

Sensitivity, specificity, pretest probability, positive and negative predictive value were calculated. For the calculations, purely screening, *i*.*e*., subjects without symptoms and without known positive households were not considered, since there is no justification to search for COVID-19 in this population.

## Results

In total, 1,757 patients were screened for COVID-19, including 1,557 and 200 patients for the retrospective and prospective analysis, respectively, and 1041 males and 716 females. In the first period, 755 patients were screened, including 413 males and 342 females. In second period, 1002 patients were screened, including 628 males and 374 females. No non-binary or non-cissexual subjects were screened.

Patients diagnosed with COVID-19 included 585 from the observational study, 94 patients from the spironolactone arm of the AndroCoV Trial (SPIRO AndroCoV-Trial), 138 patients from the dutasteride arm of the AndroCoV Trial (DUTA AndroCoV-Trial), 169 patients from the proxalutamide arm of the AndroCoV Trial (PROXA AndroCoV-Trial), and 198 patients followed apart from any of the Trials.

The positivity rates of the rtPCR-SARS-CoV-2 tests according to the number of symptoms, presence of anosmia or ageusia, and contact with a positive household for males, females, and both, in the first and second period, and altogether, are displayed in Figure 1. Positivity rates were above 60% when at least two symptoms were present, irrespective of household contact, above 80% when at least one symptom was present with known positive household contact or three or more symptoms were present without known contact, and above 95% when anosmia was present, irrespective of previous known contact with positive households, or three or more symptoms with known positive household. All patients with anosmia or ageusia and known positive household were positive for COVID-19.

**Figure 1.**
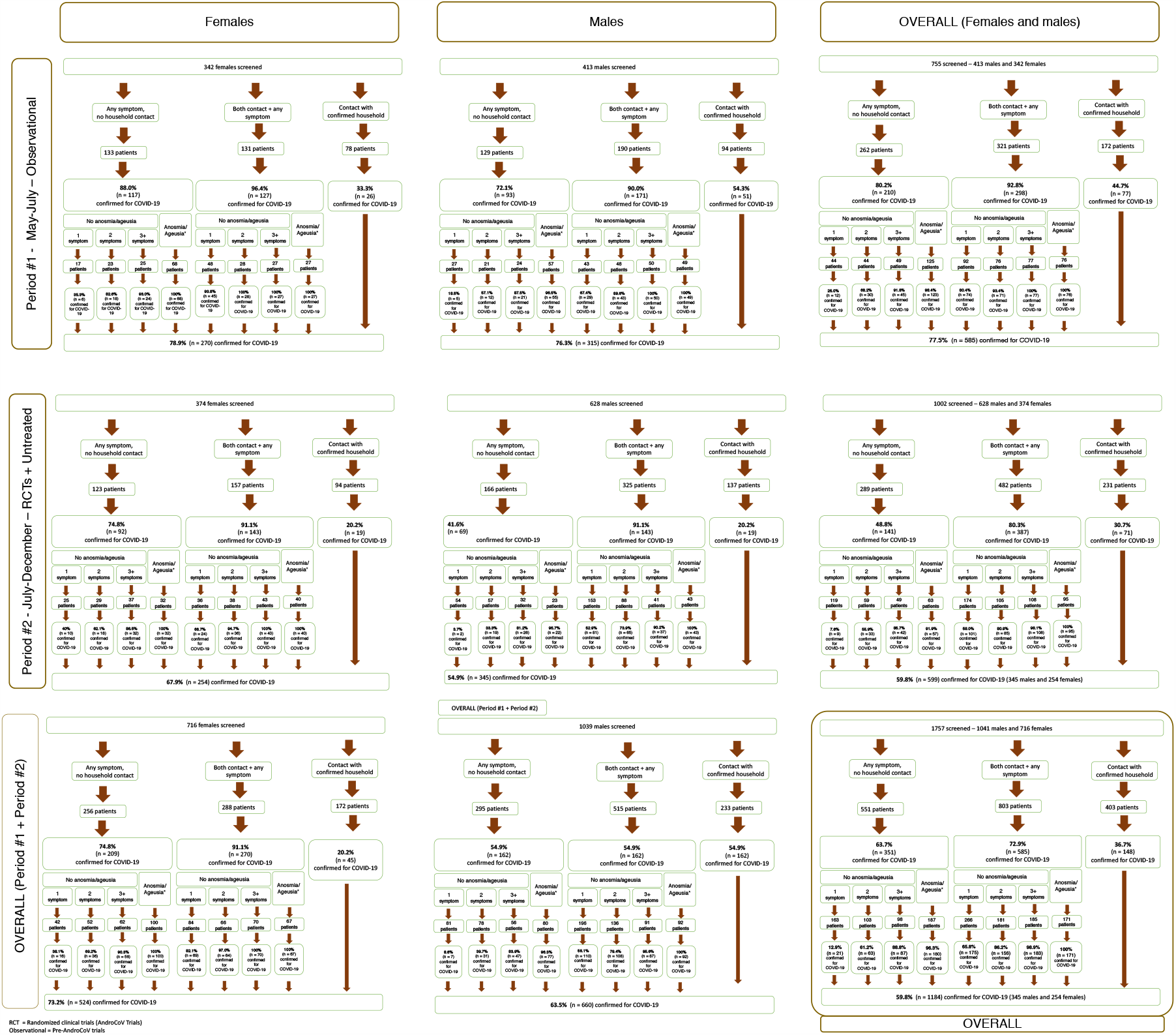
Positivity rates for rtPCR-SARS-CoV-2 according to clinical characteristics, sex, and period.

Figure 2 presents the sensitivity, specificity, PPV and NPV values to detect COVID-19 using clinical scorings in different combinations, according to the number of symptoms required or presence of anosmia when with and without known positive households. Figure 3 illustrates the tables with the number of subjects encompassed in each combination, as well as the number of true positives, true negatives, false positives and false negatives, and sensitivity, specificity, PPV, NPV, accuracy, and positive likelihood ratio. The combinations with sensitivity above 80% and accuracy above 70%, *i.e*., a pretest probability higher than the rtPCR-SARS-CoV-2, include when it is required: 1. At least one symptom is present, with or without known positive household; 2. At least two symptoms without known positive household, or with known positive household with or without symptoms; 3. At least two symptoms without known positive household or at least one symptom with known positive household; 4. At least three symptoms without known positive household or whenever there was contact with a positive household; 5. At least three symptoms without known positive household or whenever there was contact with a positive household; or 6. When anosmia or ageusia is present, with or without known positive household, or whenever there was contact with a positive household. Among these, when two or more symptoms without known contact or one or more symptoms with known contact presented the highest accuracy (80.4%).

**Figure 2.**
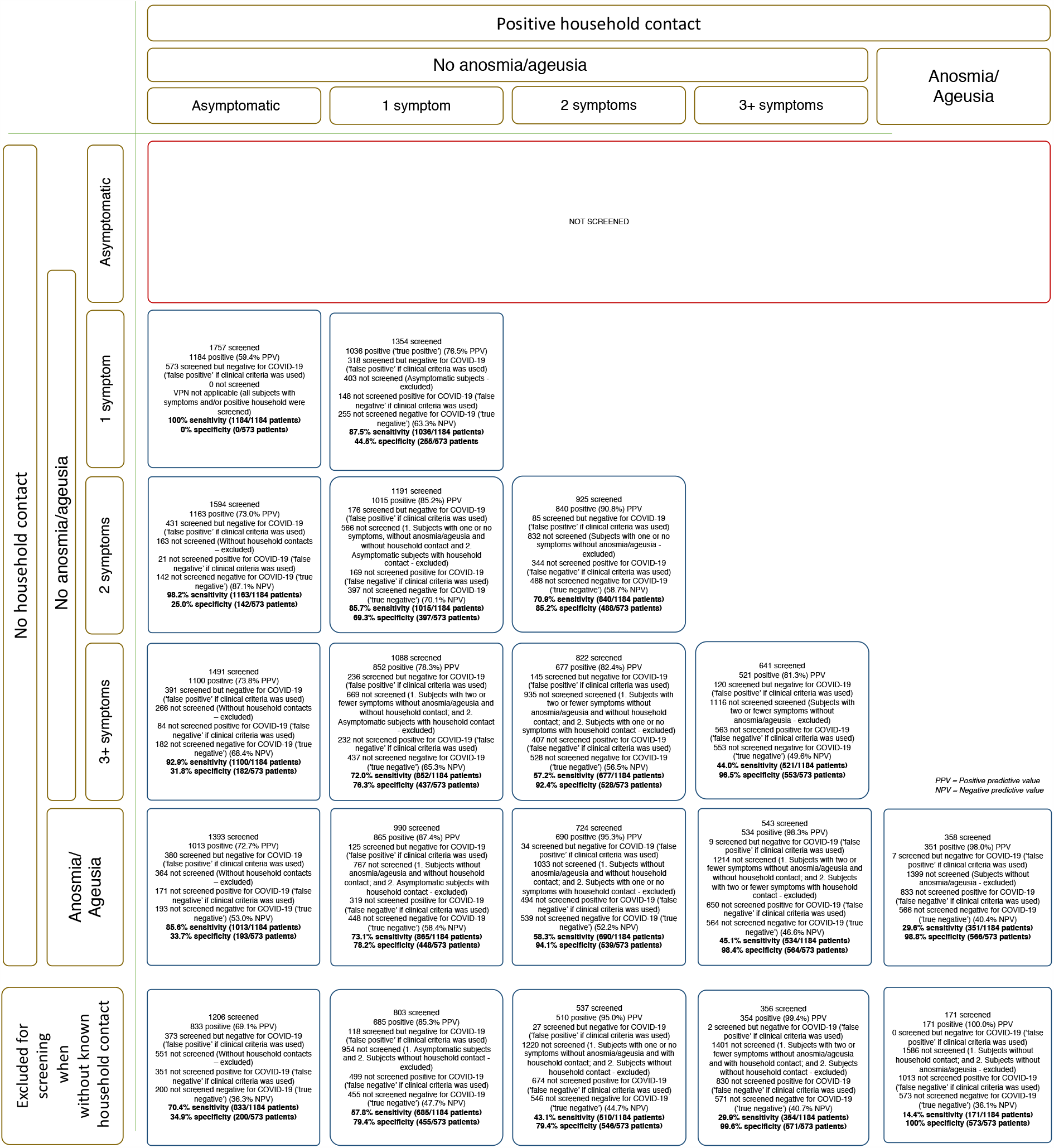
Descriptive AndroCoV Clinical Diagnostic Scoring combinations.

**Figure 3.**
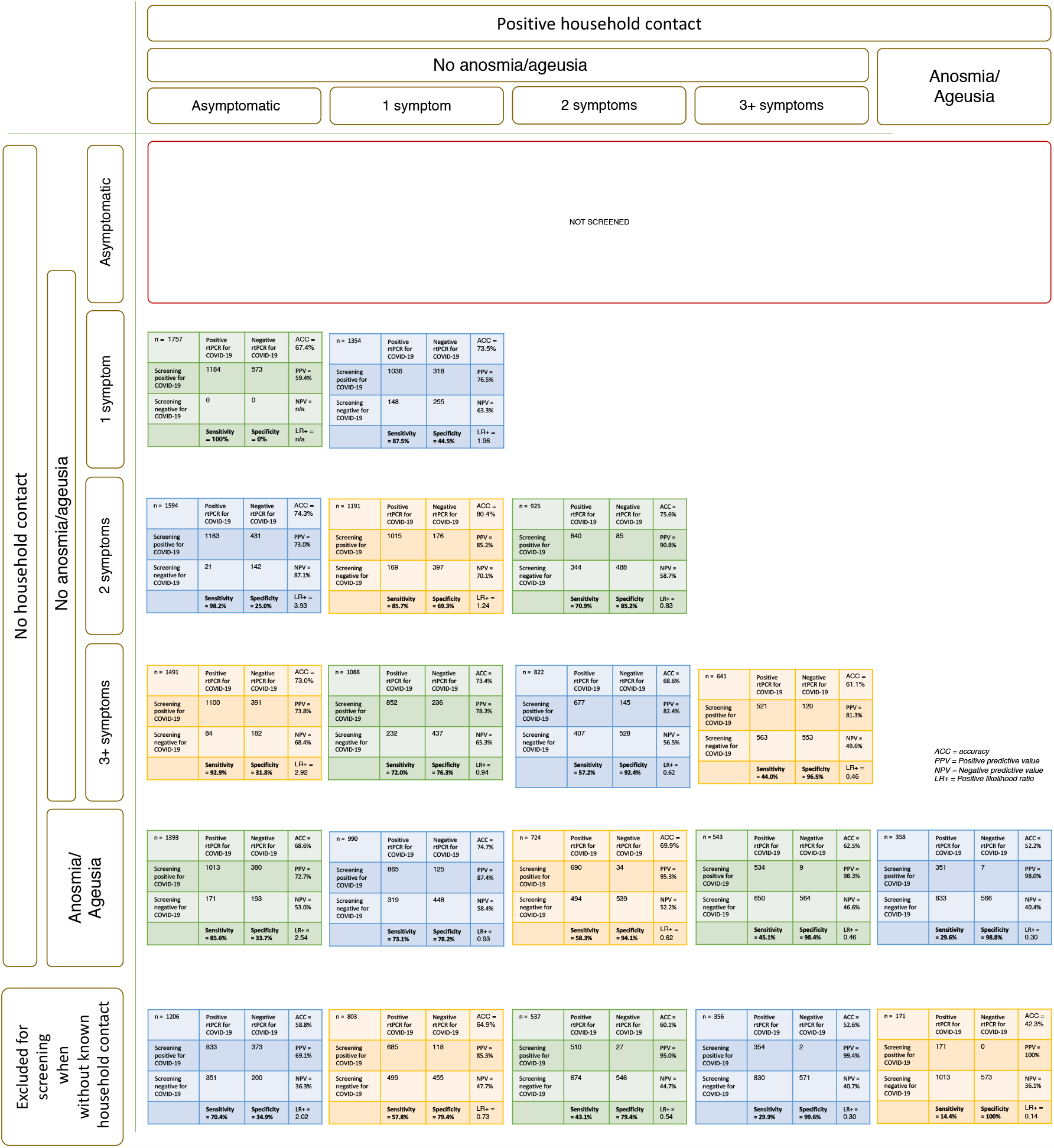
Illustrative AndroCoV Clinical Diagnostic Scoring combinations.

Figure 4 displays the recommended diagnostic management in suspected cases according to number of symptoms, presence of anosmia or ageusia, and contact with positive household. Recommendations for the management were based on the pre-test probability compared to rtPCR-SARS-CoV-2 and risk of complications from COVID-19, when delays should be avoided. In the current moment, during the COVID-19 pandemic, spread use of masks, and before vaccination reached 70% of the population, when three or more symptoms, among the ones listed, or anosmia or ageusia are present, irrespective of known positive contact, or when at least one symptom, anosmia or ageusia is present after contact with a positive household, COVID-19 can be diagnosed clinically and managed accordingly. In case two symptoms are present without known contact, rtPCR-SARS-CoV-2 should be performed, but high-risk patients should start specific therapeutics without further delays, as COVID-19 is likely present in this scenario. In case one symptom is present without contact with positive household, or when subject is asymptomatic with a positive household contact, rtPCR-SARS-CoV-2 should only be performed in high-risk patients.

**Figure 4.**
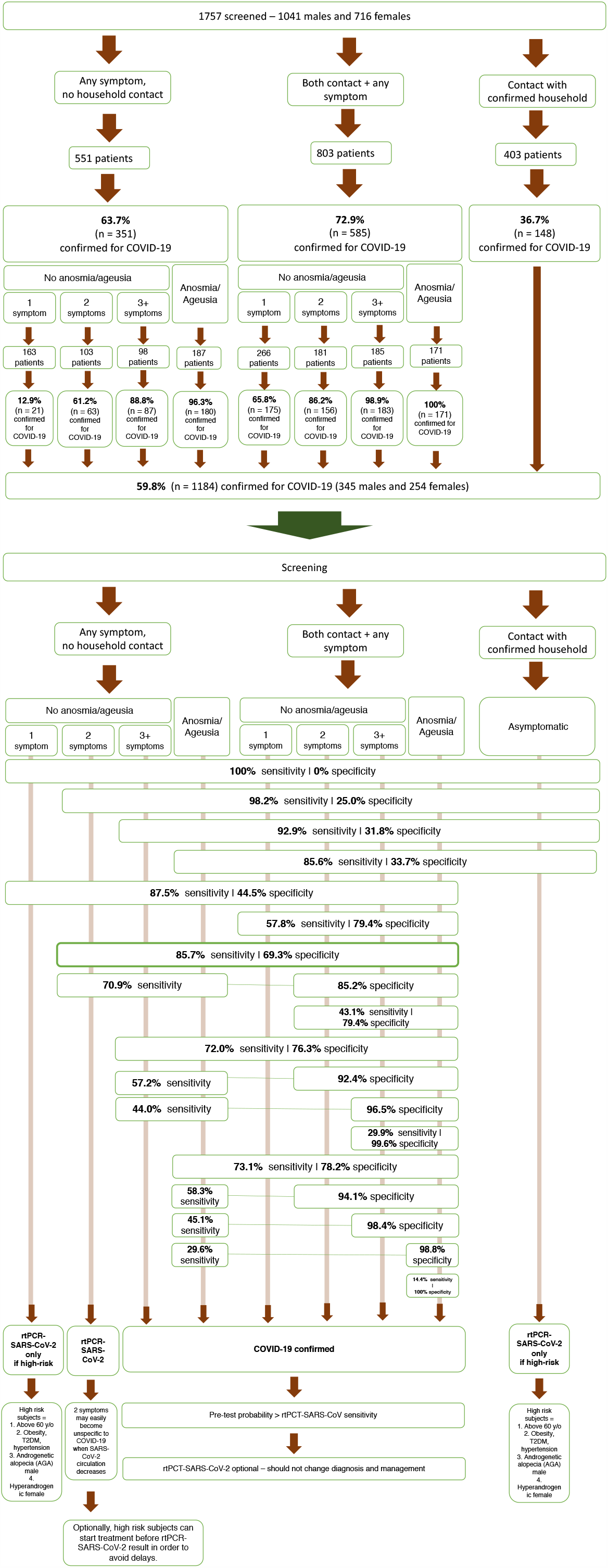
Diagnostic management for COVID-19 according to clinical characteristics and known household contact.

### Prospective follow-up

From the diagnostic management proposed in Figure 4, 200 patients were screened using the AndroCoV0derived Diagnostic Management flowchart and followed prospectively, including 169 from the PROXA Andro-CoV Trial and 29 that followed up apart from the RCTs.

Of these, 169 (84.5%) were virologically diagnosed in the first rtPCR-SARS-CoV-2, 29 (11.5%) were diagnosed in the second rtPCR-SARS-CoV-2, and two (1%) remained negative. Using two consecutive rtPCRs, the accuracy of the proposed clinical scoring combinations was 99% (Figure 5).

**Figure 5.**
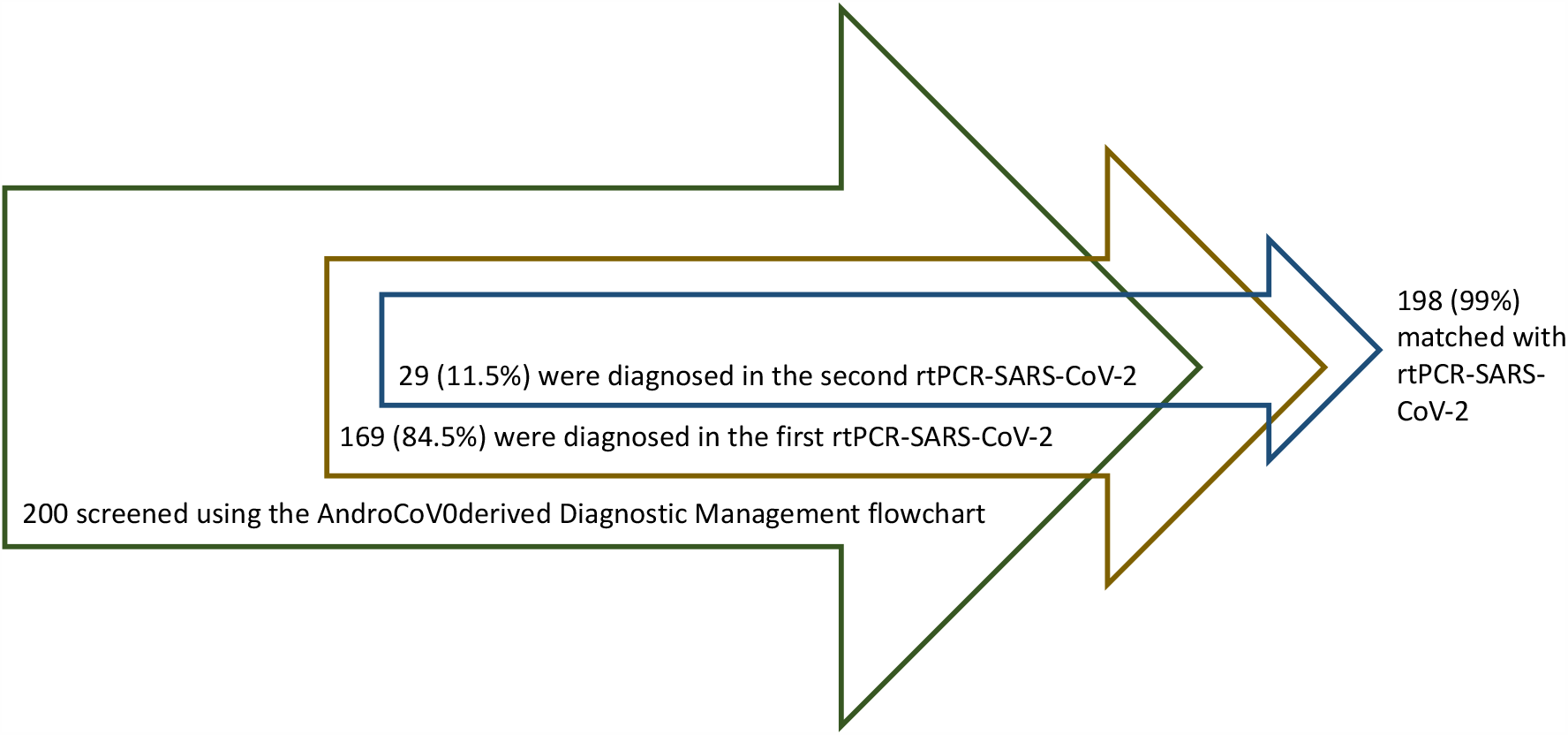
Simplified application of the presumed diagnosis of COVID-19.

### Clinical Scoring for COVID-19

From the results of the 1,757-subject cohort of the AndroCoV trials and the results presented in Figures 1 to 5, a scoring for the clinical diagnosis of COVID-19, coined as The AndroCoV Clinical Scoring for COVID-19 Diagnosis, was developed and validated, based on likelihood of a subject to present COVID-19 according to the number of symptoms, presence of anosmia, and contact with known positive household. Characteristics more specifically and critically related to COVID-19 have more points. The pointing system that best matched the most accurate clinical diagnosis is displayed in Figure 6.

**Figure 6.**
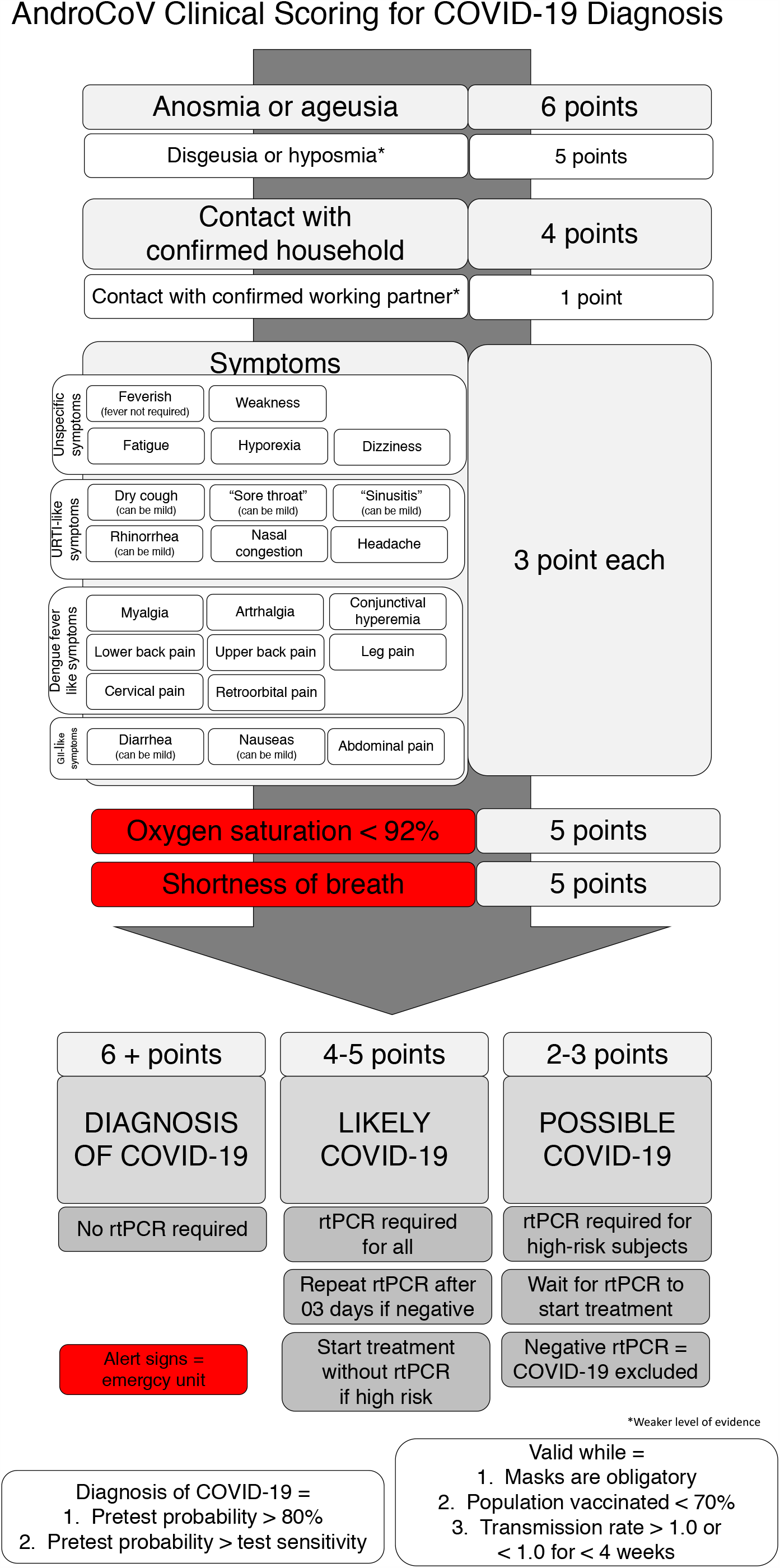
AndroCoV Clinical COVID-19 diagnosis.

**Figure 6.**
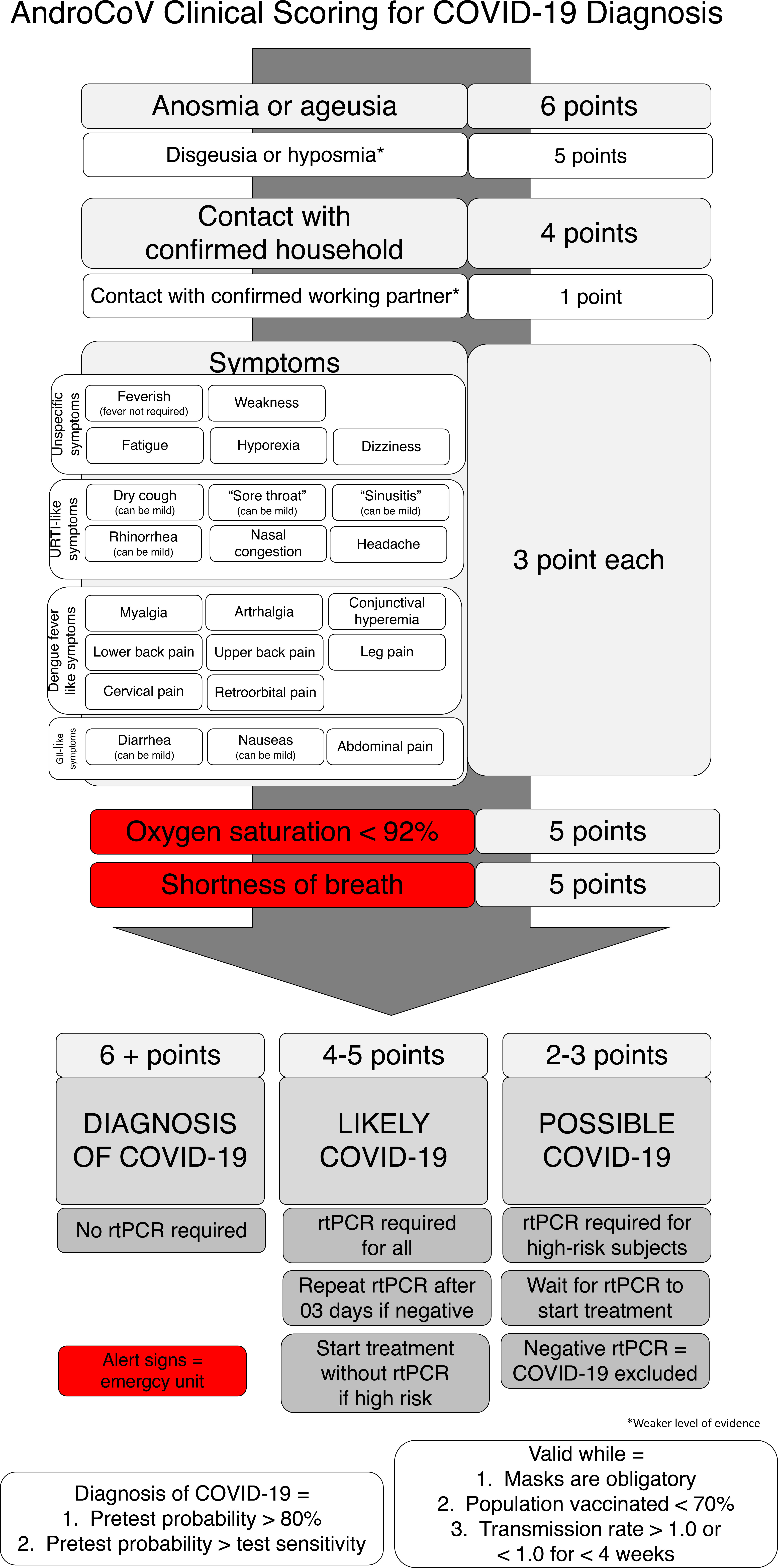
AndroCoV Clinical Scoring for COVID-19 Diagnosis.

For the clinical diagnosis of COVID-19, 6 or more points are necessary. When 6 or more points are scored, rtPCR-SARS-CoV-2 was found to be unnecessary, since the pre-test probability is higher than the rtPCR-SARS-CoV-2 sensitivity, and may lead to misdiagnosis, rather than clarification, if performed.

When between 4 and 5 points, the diagnosis of COVID-19 is likely, a rtPCR-SARS-CoV-2 is required, and if negative, a second, consecutive rtPCR-SARS-CoV-2 should be performed three days after the first test, since the sensitivity of rtPCR-SARS-CoV-2 tends to be lower in the beginning of the disease. Exceptionally, for high-risk patients, specific approaches or treatments for COVID-19 should not be delayed until a positive rtPCR-SARS-CoV-2, and should be continued independently of rtPCR-SARS-CoV-2. Three points or below represents a scenario of possible but not likely COVID-19, and rtPCR-SARS-CoV-2 is only recommended for high risk subjects.

The number of points necessary to allow the clinical diagnosis of COVID-19 was based on the likelihood of having COVID-19 when compared to the sensitivity of rtPCR-SARS-CoV-2. When the pre-test probability was higher than 80% and also higher than the rtPCR-SARS-CoV-2 sensitivity, a clinical diagnosis of COVID-19 could be determined.

The presence of anosmia or ageusia is highly specific to COVID-19 that occurs later in the first stage of the disease, and has alone a specificity of 98.8% for the COVID-19 diagnosis, irrespective of positive household contact, and should provide a more accurate COVID-19 diagnosis *per se* than rtPCR-SARS-CoV-2. For this reason, any of these two symptoms provide 06 points in the scoring, sufficient for the clinical diagnosis of COVID-19. Hyposmia and dysgeusia are highly specific as well, but may suffer interferences of other URTIs, and provide therefore 05 points. Only those with anosmia or hyposmia prior to COVID-19 should be excluded for this evaluation.

Contact with a household confirmed for COVID-19 raises the risk of COVID-19 to 50% to 60% if the positive contact was a female and 20% to 30% if the positive contact was a male, as per our analysis of the same set of subjects (2;13-15). Although the risk of transmission is approximately the double when the positive contact is a female, contact with positive female and male should not count as 04 and 02 points, respectively, because females tend to have fewer symptoms and demonstrated for specificity for the diagnosis of COVID-19 when presenting one or two symptoms. Hence, the relative importance of a positive contact is higher for females than males, which counterbalance with lower risk to be infected from a positive male, and allows that 04 points for a positive household remains precise for both male and female contact. Accordingly, even though contact with confirmed household would be more precisely counted as 04 points, the relative importance and the specificity for a positive contact for the clinical diagnosis of COVID-19 should increase with the relative decrease in SARS-CoV-2 environmental circulation compared to other microorganisms. Conversely, contact with positive working partner also raises the risk of COVID-19, although less substantially than when living with a positive contact. For this reason, a positive working partner counts as 01 point.

Except for anosmia and ageusia, since symptoms of COVID-19 are unspecific in the beginning of the disease, each symptom, not restricted to those classical ones, should count as 03 points each. For matching similar sensitivity, two symptoms is sufficient for the clinical diagnosis, since the chances of having infections other than COVID-19 are low during the prevailing circulation of SARS-CoV-2 and spread use of masks. However, the anxiety generated by the pandemic and the common inability to differentiate between previous symptom patterns and new-onset symptoms may lead to overdiagnosis of COVID-19, and this should be always considered when COVID-19 related anxiety states are detected.

As shown in the scoring system, 04 points are sufficient to start specific approaches and treatments in case of high risk patients. This is particularly important for elders since their clinical presentation may not always be as typical and may be blunted.

Finally, the presence of anosmia or ageusia, contact with a positive household and at least one symptom, or the presence of three or more symptoms irrespective of known positive contact are the three major key clinical diagnostic possibilities.

The present scoring is valid while use of masks is obligatory, population vaccinated is below 70%, and transmission rate is > 1.0 or < 1.0 for less than four weeks. Increase for 04 points for the diagnosis of COVID-19 should be considered if any of these criteria is no longer met.

### High risk patients

The determination of which subjects were at high risk for COVID-19 was based on the medical literature, and include subjects above 60 y/o, males with androgenetic alopecia (AGA), females with hyperandrogenic states, and those presenting metabolic-related conditions, including obesity, diabetes mellitus, and hypertension (16).

### Follow-up

Subjects that do not fulfill criteria for COVID-19 should be reassessed for clinical symptoms and contact on a daily basis in the following three days, since new symptoms should appear on those who present actual COVID-19, which will raise the score in the following days and allow the clinical diagnosis of COVID-19, timely for appropriate management, before any complication. Patients with 4 or 5 points should be particularly reassessed, once COVID-19 progresses to more severe states quickly. These patients should be clinically reassessed even with a negative rtPCR-SARS-CoV-2 since the sensitivity of this test is lower than 80% to 90%, and tends to be lower in the early days of the disease.

## Discussion

Clinical diagnosis of COVID-19 finds multiple advantages over the need of a rtPCR-SARS-CoV-2 test, including reduction of costs, since rtPCR-SARS-CoV-2 has a relatively high cost, prevention of further delays in therapeutical approaches for COVID-19, and few severe harms compared to multiple potential benefits of early and ‘overdiagnosis’, in comparison to ‘underdiagnosis’, of COVID-19.

With the present thorough analysis of 1,757 subjects suspected for COVID-19, we found sufficient substantiation to recommend against a mandatory rtPCR-SARS-CoV-2 for the diagnosis of COVID-19 in highly suspected subjects. This should reduce the screening costs and the inequity caused by the lack of wide access to rtPCR-SARS-CoV-2. For patients clinically diagnosed for COVID-19 through our clinical scoring system, rtPCR-SARS-CoV-2 should be avoided because clinical diagnosis has demonstrated higher accuracy than virological one, at least when compared to commercially available rtPCR-SARS-CoV-2 kit tests. In case of a negative rtPCR-SARS-CoV-2, because of its overwhelming risk of being a false negative result, clinical diagnosis, rather than test result, must be considered.

To overcome false negative rtPCR-SARS-CoV-2 that may lead to loss of timely detection of subjects developing severe COVID-19, we proposed a for moderately suspected patients, a second consecutive rtPCR-SARS-CoV-2 to be repeated between 24 and 72 hours after the first test, since more than 80% of those with a first negative rtPCR-SARS-CoV-2 showed a positive result when performed again.

A clinical, early diagnosis of COVID-19 is particularly important for subjects at higher risk to develop severe COVID-19. Elders, for instance, may present even lower sensitivity for a rtPCR-SARS-CoV-2 test. In addition, their clinical presentation may not always be as typical as the already unspecific symptoms found in COVID-19. This population could be particularly benefited from the clinical diagnosis of COVID-19 in order to prevent the development of more severe states.

Additional interesting findings were unveiled by the present analysis. Only 1 in every 7 subjects with COVID-19 had anosmia or ageusia with known household contact. This means that for every 7 patients with COVID-19, 6 will not present anosmia and known contact with positive household. This finding finds importance in the policies for COVID-19 diagnosis.

The unique characteristics of the pandemic and the peculiarities of the virus does not allow an undisputed method of establishing clinical criteria. However, by assessing subjects with any type of sign to suspect for COVID-19 – symptoms, confirmed contact, or both – this clinical diagnostic tool represented a virtually 100% sensitive flowchart. The only non-encompassed group that could miss sensitivity were asymptomatic patients without known confirmed contacts. However, this population is highly unlikely affected, and should be the least priority when tested.

Shortness of breath with oxygen saturation > 94% is more likely due to anxiety induced by COVID-19 than the disease *per se*. Oppositely, the ‘happy hypoxia’ shows that shortness of breath due to COVID-19 only occurs when oxygen saturation are overtly low. However, since the present clinical diagnosis aims to counteract with the prevailing inertia that led to an excessive number of deaths due to COVID-19, we recommend for the investigation of shortness of breath, regardless of oxygen saturation.

Similarly, although the number of symptoms alone can lead to a large number of false positive COVID-19 diagnosis, the counterbalance for the highly specific but not sensitive rtPCR-SARS-CoV-2 in the current context of the pandemics, when high sensitivity must be targeted.

The prevalence of anosmia and ageusia was lower than our data of the trials because some of the subjects positive for COVID-19 developed these symptoms after the diagnosis.

### Recommendations based on the findings

1. While tests are not extremely sensitive and pretests are high, the employment of rtPCT-SARS-CoV-2 as the sole diagnostic method for patients with pretest probability above 80% should be considered a misuse of the test.
2. Since sensitivity also varies according to the viral load, clinical diagnosis should be preferred over virological methods during the pandemics.
3. We recommend against the use of rtPCR-SARS-CoV-2 if our proposed scores indicate the diagnosis of COVID-19.
4. While determining the exact threshold of the pretest, above which a biochemical test becomes unnecessary, although challenging, is questionable in the current context, because factors used to determine the thresholds should not only depend on the test sensitivity and posttest probability, but also on the numbers and consequences of missing COVID-19 diagnosis.
5. Whenever the chances of corresponding positive test is above its sensitivity, we considered the clinical diagnosis as presumed.
6. Scoring points may be adapted according to region-specific clinical presentation, transmission rates, and potential viral mutations
7. The present clinical scoring for clinical diagnosis of COVID-19 should only be valid when transmission rate is above 1.0 or below 1.0 for less than 4 weeks and masks are widely used.
8. The present score should be reassessed after 70% of population has been vaccinated.
9. We recommend for the reassessment of drugs with potential antiviral activity when using the present, actual early COVID-19 diagnosis, since they may present effectiveness if used early, unlike the lack of results when administrated later in the disease.
10. While SARS-CoV-2 remains as the prevailing circulating virus, masks effectively block bacterial infections, and rtPCR-SARS-CoV-2 still under-detect COVID-19, the use of a score for clinical diagnosis of COVID-19 should be considered as the first line diagnostic tool.

## Limitations

While we matched rtPCR results with clinical aspects to determine the pretest probability, this determination of pretest probability is imprecise due to the still challenging and largely unclear understanding of the COVID-19 transmission patterns. The present scoring system was based on SARS-CoV-2 transmission and clinical characteristics of a specific region, and may not precisely reflect the patterns present in other regions.

## Conclusion

The AndroCoV Clinical Scoring for COVID-19 diagnosis was demonstrated to be a feasible, fast, inexpensive and sensitive diagnostic tool for a clinical diagnosis of COVID-19, that avoids delays and missed diagnosis, and should be recommended as a first-line option for COVID-19 diagnosis for public health policies.

## Data Availability

Full raw data of the observational AndroCoV trials are available at a public repository, in the following electronic address: https://osf.io/cm4f8/, that has been made public available. Data from the AndroCoV RCTs and for the present study are available under request to.

## Acknowledgments

We acknowledge the full Corpometria Institute and Applied Biology Inc teams, who actively helped to screen and enroll patients, and provided full support to patients during their period of the proposed treatment for COVID-19 We also acknowledge Laboratório Exame, from DASA Diagnósticos da América (Brasília, DF and São Paulo, SP, Brazil), for their efforts to make the present study financially feasible to be supported by Corpometria Institute and Applied Biology Inc, and for providing physical space for the research with no cost, and EMS Pharmacological Industry (Campinas, SP, Brazil).

## Funding statement

The funding of present study was fully supported by Corpometria Institute (Brasilia, DF, Brazil) and Applied Biology Inc (Irvine, CA, USA).

## Conflict of interest statement

Authors declare no conflict of interest with any of the pharmacological interventions proposed by the present study.

## Ethics approval statement

The present study was approved by the Institutional Review Board (IRB) of the Ethics Committee of the National Board of Ethics Committee of the Ministry of Health, Brazil (CEP/CONEP: Parecer 4.173.074 / CAAE: 34110420.2.0000.0008), and is registered at ClinicalTrials.gov (Identifier: NCT04446429. Available at clinicaltrials.gov (https://clinicaltrials.gov/ct2/show/NCT04446429?term=NCT04446429&draw=2&rank=1).

## Patient consent statement

Patients included for the present analysis provided a written consent exactly as approved by the Institutional Review Board (IRB) of the Ethics Committee of the National Board of Ethics Committee of the Ministry of Health, Brazil (CEP/CONEP: Parecer 4.173.074 / CAAE: 34110420.2.0000.0008), alongside with the currently ongoing randomized clinical trial (RCT) registered at ClinicalTrials.gov (Identifier: NCT04446429. Available at clinicaltrials.gov (https://clinicaltrials.gov/ct2/show/NCT04446429?term=NCT04446429&draw=2&rank=1).

## Authorship

Authors FAC, AG, CGW and JM contributed equally for the present study. FAC, AG, CGW and JM designed the study. FAC enrolled, provided medical assistance to all patients and compiled all data, while FAC, AG, CGW and JM contributed for the analysis of the data and development of the manuscript in its current format.

## List of abbreviations

PPV: positive predictive value
NPV: negative predictive value
rtPCR: real-time polymerase chain reaction
COVID-19: infection caused by the Severe Acute Respiratory Syndrome Coronavirus 2 (SARS-CoV-2)
SARS-CoV-2: Severe Acute Respiratory Syndrome Coronavirus 2 (SARS-CoV-2)

## Notes

### Competing Interest Statement

The authors have declared no competing interest.

### Clinical Trial

NCT04446429

### Author Declarations

The study was approved by an ethics committee and registered with clinicaltrials.gov (NCT04446429), and also approved by Brazilian National Ethics Committee (Approval number 4.173.074; CAAE 34110420.2.0000.0008; Comite de Etica em Pesquisa (CEP) of the Comite Nacional de Etica em Pesquisa (CONEP) of the Ministry of Health (MS)) (CEP/CONEP/MS).

